# Cytokine Profiling of Children, Adolescents, and Young Adults Newly Diagnosed with Sarcomas Demonstrates a Role for IL-1β in Osteosarcoma Metastasis

**DOI:** 10.1101/2025.04.05.25325205

**Authors:** Laurel Kastner, William Kandalaft, Aakash Mahant Mahant, Jessica Crimella, Sydney Hakim, Xiao Peng, Michael S. Isakoff, Masanori Hayashi, David M. Loeb

**Affiliations:** Department of Pediatrics, Albert Einstein College of Medicine, Bronx, NY 10461; Department of Developmental and Molecular Biology, Albert Einstein College of Medicine, Bronx, NY 10461; Moffitt Cancer Center, Tampa, FL 33612; Department of Pediatrics-Hematology/Oncology and Bone Marrow Transplantation, University of Colorado, Aurora, Colorado 80045; University of Connecticut School of Medicine, Hartford, CT 06106; Montefiore Einstein Comprehensive Cancer Center, Albert Einstein College of Medicine, Bronx, NY 10461; Cancer Dormancy Institute, Albert Einstein College of Medicine, Bronx, NY 10461; Marilyn and Stanley M. Katz Institute for Immunotherapy for Cancer and Inflammatory Disorders, Albert Einstein College of Medicine, Bronx, NY 10461

## Abstract

**Background:** Sarcomas are a heterogeneous group of mesenchymal tumors frequently diagnosed in pediatric and young adult patients. These tumors respond poorly to conventional immunotherapy, though the precise reason for this is not known. We sought to characterize the systemic immune response to sarcomas by measuring the levels of circulating cytokines in the plasma of sarcoma patients, testing the hypothesis that the natures of a patient’s immune response to their tumor directly affects outcome.

**Methods:** Plasma was collected from newly diagnosed, treatment-naive pediatric sarcoma patients participating in an ongoing clinical trial, MCC20320. A panel of 18 cytokines was selected and cytokine levels were measured using the Luminex platform. Cytokine levels were analyzed based on clinicopathological parameters such as gender, age, stage, and survival.

**Results:** We found that the cytokine profile in patients newly diagnosed with sarcoma is distinct from healthy controls, but different sarcomas were not distinguishable. Patients with osteosarcoma who had elevated levels of multiple cytokines had inferior overall survival compared to those with fewer or no elevated levels. Similarly, elevated levels of individual cytokines and chemokines, including IL-24, CXCL5, and CXCL10, were associated with inferior event-free or overall survival in patients with osteosarcoma. Perhaps most significantly, elevated IL-1β at diagnosis was associated with metastatic presentation and inferior event-free survival in patients with osteosarcoma.

**Conclusion:** These findings suggest that pediatric sarcoma patients mount a systemic immune response that may affect event-free or overall survival. IL-1β in particular may be a valuable target for immunotherapy for osteosarcoma patients.

**Statement of Translational Relevance:** We report the results of a prospective study profiling cytokine levels in the serum of newly diagnosed, treatment-naïve children and young adults with sarcomas. We found elevated levels of several pro-inflammatory cytokines in the serum of these patients. Elevated levels of several of these, including IL-24, CXCL5, and CXCL10, were associated with inferior event-free or overall survival in patients with osteosarcoma. We also found that elevated IL-1β at diagnosis was associated with metastatic presentation and inferior event-free survival in patients with osteosarcoma. In the context of previously published preclinical work demonstrating that blocking IL-1 signaling can inhibit osteosarcoma metastasis, our work supports development of a clinical trial testing this concept in patients with osteosarcoma.

## Introduction

Immune activity in the tumor microenvironment (TME) is widely appreciated for its ability to affect oncogenic disease establishment and progression through numerous mechanisms, including modulation of tumor proliferation, immune cell response to tumor cells, angiogenesis, tumor cell metabolism, and dissemination of tumor cells^1,2^. Immune and non-immune cells in the TME establish a complex, dynamic constellation of signaling crosstalk that evolves along with the tumor. Cytokine and chemokine measurements are increasingly obtained as surrogate measurements of immune and inflammatory signaling activities because of the diversity of cell-intrinsic, innate, and adaptive immune processes that produce them; however, not all important immune signaling processes are equally well-represented by this strategy, as has been shown for type I interferon activation^3^. Moreover, interpreting the role played by any individual signal is complicated because its impact is always contextual and never isolated. The level at any particular timepoint is a composite of reactive and inductive contributions from many cell types and even potentially opposing processes, varying based on the identity and nature of cells within the TME, the level and duration of expression and action, and the combined impact with other signals^4,5^. This complexity necessitates studies evaluating diverse circumstances of cytokine activity in different disease endotypes and environments. It is difficult to extrapolate from one tumor type to another because each type of tumor elicits a unique immunologic response. In addition, profiling multiple cytokines simultaneously is required to fully understand tumor immune responses. Many cytokines can circulate systemically, producing dramatic effects even at a distance from the tumor. Despite this complexity, studies have found that characterizing circulating cytokines can predict disease outcomes in some cases^6–8^.

With a unique mesenchymal lineage, sarcomas are rare and less well-studied than other solid tumors in adults; however, they are more common in children, adolescents, and young adults^9^. Because of their heterogeneous nature and relative rarity, little is known about the immune response to these tumors. In general, sarcomas are relatively unresponsive to immunotherapy, but the mechanisms underlying this relative lack of response are poorly understood^10,11^.

In this study, we characterized the cytokine profiles of children, adolescents, and young adults newly diagnosed with Ewing sarcoma, osteosarcoma, rhabdomyosarcoma, and other subtypes. The goal of this work was to test the hypothesis that the nature of the immunologic response to a tumor affects patient outcome. We simultaneously quantified serum levels of 13 pro-tumorigenic^12–21^ and 5 anti-tumorigenic^22–26^ cytokines selected based on evidence suggesting they played a role in one or more sarcomas^8,13,19,24,25,27–40^. We found that specific cytokine levels correlate with key features of disease outcome. Most significantly, we found that patients with osteosarcoma who have high levels of plasma IL-1β at diagnosis are more likely to have metastatic disease at presentation and have a shorter event-free survival (EFS) than those with lower levels of this cytokine.

## Methods

### Patients

Research subjects were enrolled in an ongoing multi-center clinical trial, MCC20320, a study evaluating blood-based biomarkers of disease response and prognosis in children, adolescents, and young adults with sarcomas. This multi-center clinical trial was approved by Advarra, and was conducted in accordance with the Ethical Principles and Guidelines for the Protection of Human Subjects of Research. Newly-diagnosed patients less than or equal to 40 years old and weighing over 8 kg were enrolled at participating centers. Cohorts were defined as follows: Cohort 1: Ewing sarcoma patients with confirmed or suspected diagnosis of Ewing sarcoma (ES) with a t(11;22) EWS-FLI1 or t(21;22) EWS-ERG translocation; Cohort 2: fusion-positive rhabdomyosarcoma (FP-RMS) patients with confirmed or suspected diagnosis of a rhabdomyosarcoma with a t(2;13) PAX3-FOXO1 or t(1;13) PAX7-FOXO1 translocation; Cohort 3: fusion negative rhabdomyosarcoma (FN-RMS) with a confirmed or suspected diagnosis of a rhabdomyosarcoma without a t(2;13) PAX3-FOXO1 or t(1;13) PAX7-FOXO1 translocation; Cohort 4: confirmed patients with osteosarcoma; Cohort 5: other translocation-driven sarcomas not included in the other 4 cohorts. Plasma from clinical waste of presumed healthy individuals was used for healthy controls. Study accrual opened in 2019 and is ongoing, but this report focuses on subjects with Ewing sarcoma and osteosarcoma, the cohorts with sufficient subject enrollment for meaningful analysis. Only treatment-naive samples drawn at the time of diagnosis were evaluated.

### Plasma Extraction

Blood was collected at enrollment in a Streck Cell-free DNA BCT® tube and shipped to the University of Colorado Sarcoma Research Lab in ATS1 temperature-regulated containers (Akuratemp). Plasma was isolated by double centrifugation and frozen at −80 °C until use, as previously described by Hayashi *et al*.^41^. Samples were aliquoted and frozen at −80 °C and underwent no more than two freeze/thaw cycles.

### Luminex Assays

All samples were measured in technical duplicates and averaged for a final concentration. IL-1β, IL-4, IL-6, IL-8, IL-10, IL-12(p40), IL-15, IL-27, and CXCL10 were measured using kit HCYTA-60K-10. CCL21, CXCL5, IL-16, IL-24, and CXCL12 were measured using kit HCYTB-60K-06. MIF, TGF-β1, and IGF-1 were measured using HSP1MAG-63K-01, TGFBMAG-64K-01, and HIGFMAG-52K-01, respectively, all obtained from Sigma Millipore. All kits were used according to the manufacturer’s instructions. Additional dilution was required to quantify IGF-1 (1:120), CXCL4 (1:20,000), and TGF-β1 (1:270). Data were acquired on a Luminex Magpix XMAP Multiplex Reader (Luminex Technologies, RRID:SCR_023348) and analyzed using Belysa Immunoassay Curve Fitting Software (Millipore Sigma). Interpolated concentrations of all analytes were log transformed before analysis to reduce non-normal skew and inter-assay variability.

### Statistical analysis

Healthy patients and sarcoma cohorts were analyzed using Singular Value Decomposition (SVD) with imputation to calculate principal component analysis (PCA) with unit variance scaling applied using ClustVis software (RRID:SCR_017133)^42^. Prediction ellipses were formed with a 95% confidence interval. ClustVis was also used to generate a heatmap of cytokine levels using unsupervised clustering on columns and rows with correlation distance and complete linkage. Based on hierarchies and visual inspection, endotypes were split into three groups, which were further analyzed for overall survival and event free survival by Log-rank (Mantel-Cox) test. This and all remaining statistical tests were performed using GraphPad Prism (RRID:SCR_002798). All analytes in osteosarcoma and Ewing sarcoma samples were also analyzed using a Spearman r correlation matrix. Receiver operating characteristics (ROC) curves and area under the curve (AUC) was calculated for osteosarcoma and ES cohorts for analytes that were significantly altered compared to healthy controls.

Individual analyte concentrations in each cohort were compared to healthy controls using a Mann-Whitney test. Further analysis was performed on groups that had sufficient sample size, including Ewing sarcoma, osteosarcoma, and all patients not separated by cohort, referred to as “mixed cohorts”. To explore the effect of multiple elevated or depressed cytokines in conjunction, patient outcomes to date (responsive, refractory, or relapsed disease) were distributed by number of elevated or depressed analytes. Cytokines were considered elevated if plasma concentration was greater than the 95^th^ percentile of the healthy cohort. Depressed levels were below the 5^th^ percentile. Additionally, patients were grouped as 0-3 and ≥4 elevated cytokines or 0 and ≥1 depressed cytokines. Based on these groupings, event free survival (EFS) and overall survival (OS) were analyzed with Kaplan-Meier curves with an 18-month endpoint, and hazard ratios were generated using a Log-rank (Mantel-Cox) test. EFS and OS were also determined for high and low cytokine levels, with high defined as anything at or above 75^th^ percentile. Further analysis was performed comparing adult (age ≥22) and pediatric (age ≤ 21) and localized or metastatic stage of bone sarcomas at diagnosis by Mann-Whitney test. Since IL-1β had compelling results, we also used Fisher’s exact test to determine if there was a difference in the number of patients with metastatic or localized disease in the high or low IL-1β groups.

## Results

A total of 123 sarcoma patients and 17 healthy subjects were included in this analysis, and they are presented in Supplementary Table 1. Median cytokine values for each cohort and their comparison to healthy controls can be found in Supplemental Table S1. When analyzed globally through PCA, all five sarcoma cohorts had overlapping clusters but were distinguishable from healthy controls (Figure 1A). Similarly, when comparing patients’ cytokine levels through unsupervised hierarchical clustering, healthy controls clustered together. In contrast, specific sarcoma cohorts were not segregated (Figure 1B). Although patients with the same diagnosis did not cluster together in this analysis, we were able to group patients into three endotypes (Figure 1B). Group 1 was distinguished by an increase in CXCL5, CXCL12, and/or MIF. Group 3 had increased levels of IL-6, IL-8, IL-4, IL-1β, IL-15, IL-10, IL-12 p40, and/or IL-24 with relatively low levels of CCL21, CXCL10, and IGF-1. Group 2 had few distinguishing features besides a slight increase in CXCL10 in approximately a third of patients. Although endotype grouping did not have statistically significant impact on survival, there were trends toward significance that might have been statistically significant with larger cohorts or longer follow-up (Figure 1C). For example, patients with osteosarcoma in Endotype 1 had the worst EFS (p=0.15), and looking at all enrolled subjects, those with Endotype 1 had the worst OS (p=0.15).

**Figure 1.**
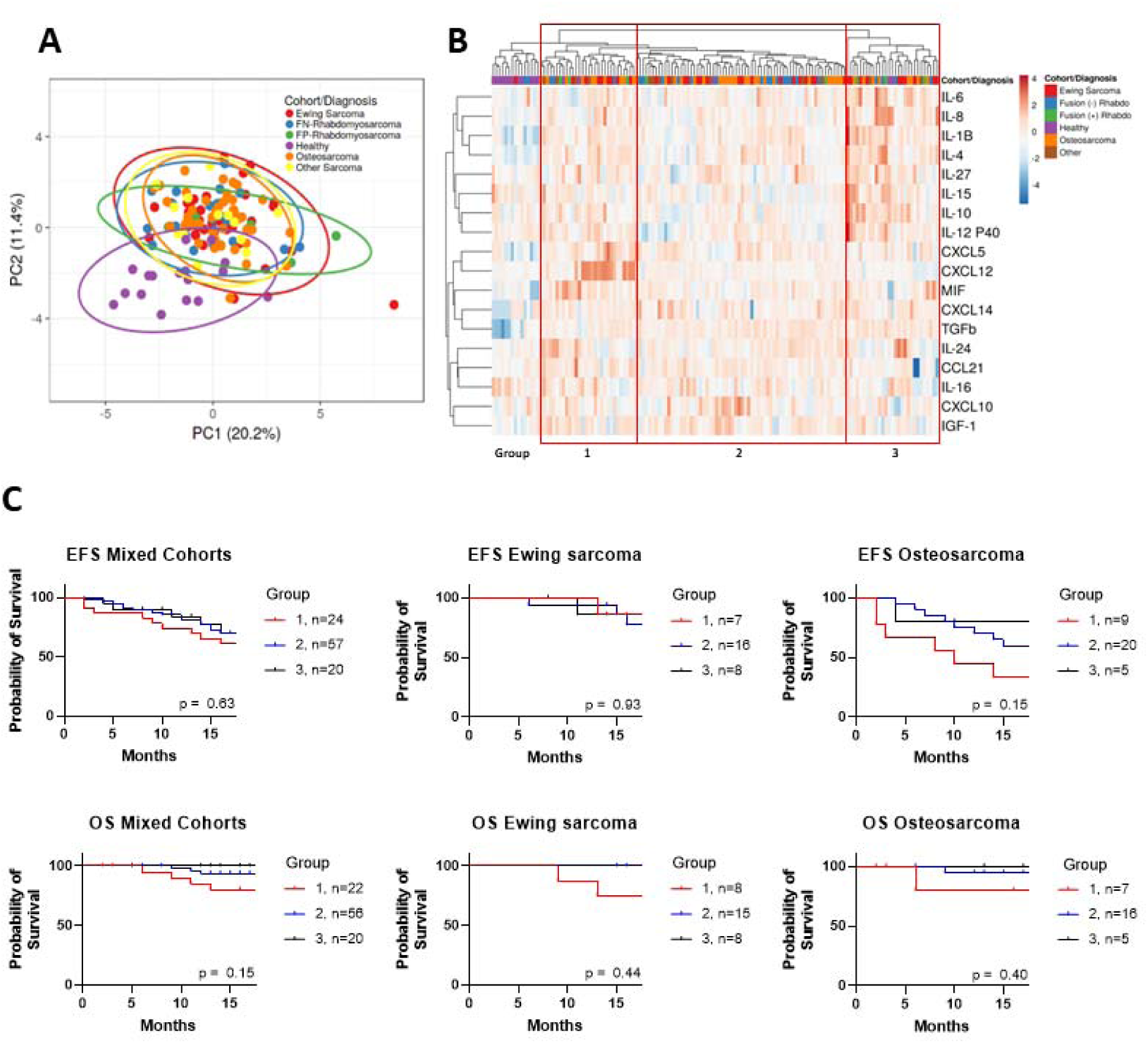
Unsupervised clustering of sarcomas compared to healthy controls. A) SVD imputation to calculate principal component analysis with unit variance scaling applied B) heat map on cytokine expression with cohort type; unit variance scaling is applied to rows. Imputation is used for missing value estimation. Both rows and columns are clustered using correlation distance and complete linkage. Red squares denote endotype grouping. C) Overall Survival (OS) and event free survival (EFS) Kaplan-Meier curves for different endotype groups. P = NS determined by Log Rank test

A Spearman correlation matrix was generated for osteosarcoma and Ewing sarcoma samples, which demonstrated several significant correlations, suggesting biologically meaningful relationships (Supplementary Figure 1). Osteosarcoma had 25 significant relationships, notably including IL-4/IL-1β (r²=0.58, p = 0.000048), IL-12 p40/IL-4 (r²=0.47, p = 0.0015), CXCL5/IL-27 (r²=-0.48, p = 0.001), CXCL14/IL-15 (r²=0.66, p=0.0000021). In Ewing sarcoma, there were 21 significant relationships including IL-4/IL-1β (r² = 0.55, p=0.00048), IL-4/IL-8 (r²=0.68, p=0.0000047), CXCL14/IL-15 (r²=0.61, p=0.00013), CXCL5/CXCL12 (r²=0.64, p=0.000030).

We next sought to determine if having multiple elevated cytokines had an impact on disease progression. Defining “elevated” as the upper 5% of the healthy cohort’s cytokine levels, 44% of Ewing sarcoma and 55% of osteosarcoma patients had four or more cytokines elevated in tandem (Figure 2A). Poor outcomes, like relapse and refractory disease, were more common among patients that had more elevated analytes in total (Figure 2B). These patients also had worse OS in osteosarcoma but not Ewing sarcoma (Figure 2C: Osteosarcoma p=0.043; Ewing sarcoma p=0.65); however, the impact on EFS did not reach statistical significance for either tumor type (Ewing sarcoma p=0.79; osteosarcoma p=0.55).

**Figure 2.**
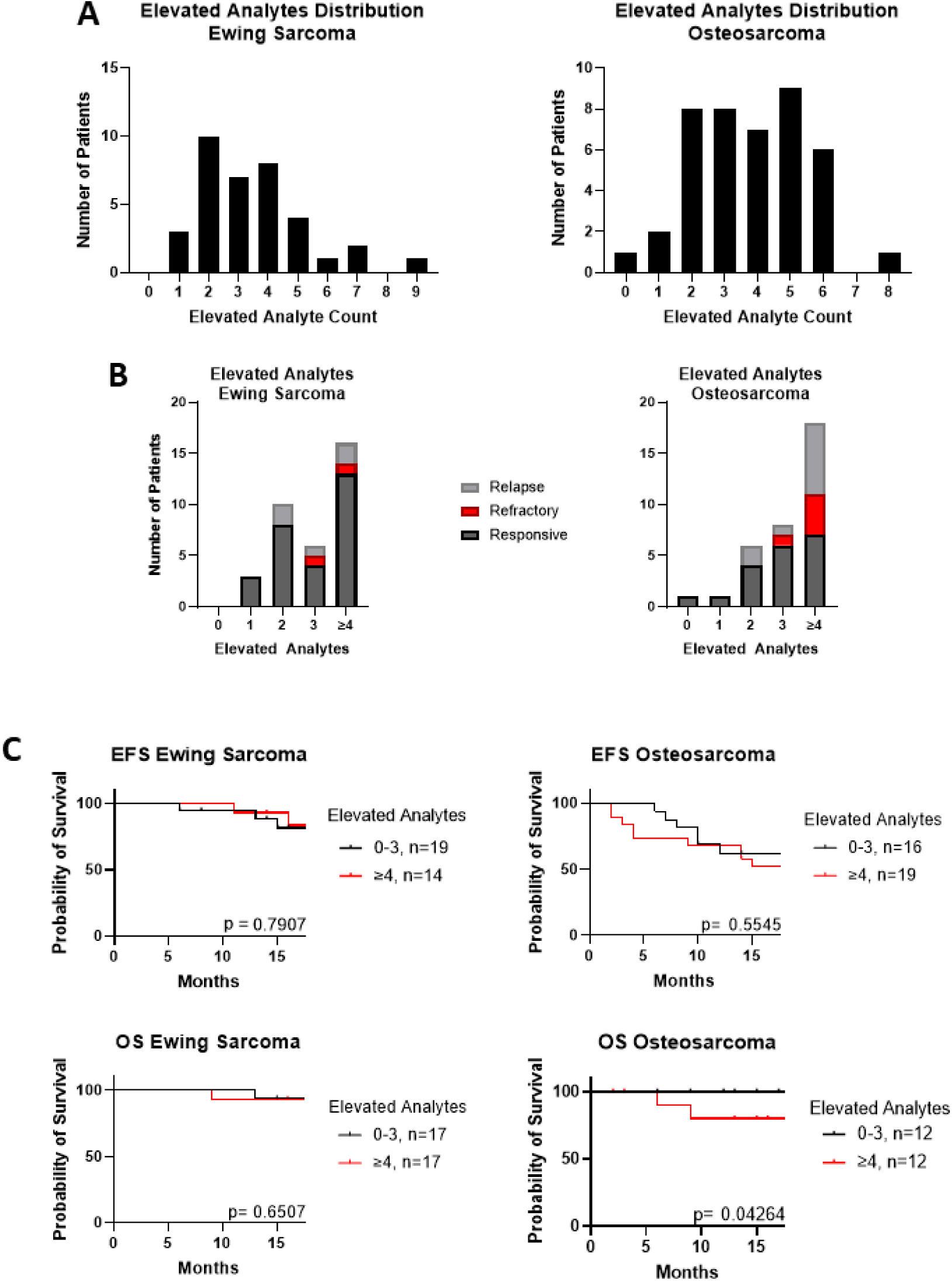
Comparison of outcome based on total number of elevated analytes, defined by top 5% of the healthy cohort’s mean. A) Distribution of osteosarcoma and Ewing sarcoma patients by their number of elevated cytokines. B) Clinical outcomes for patients separated by how many elevated cytokines they have. C) Event free survival (EFS) and overall survival (OS) curves for Ewing sarcoma and osteosarcoma patients with 0-3, or ≥4 cytokines elevated.

A complementary result was seen in patients with multiple depressed analytes. Defining “depressed” as the lower 5% of the cytokine level in healthy controls, we found that although only 6% of Ewing sarcoma and 2% of osteosarcoma patients had four or more cytokines depressed in total (Figure 3A), patients with poor outcomes had fewer depressed analytes in total (Figure 3B). Similarly, having fewer depressed cytokines correlated with slightly improved EFS, which approached significance for osteosarcoma (Figure 3C: Ewing sarcoma, p=0.52; Osteosarcoma p=0.082). The effect on OS was not statistically significant, probably because of small sample size (Ewing sarcoma p=0.75; Osteosarcoma p=0.69).

**Figure 3.**
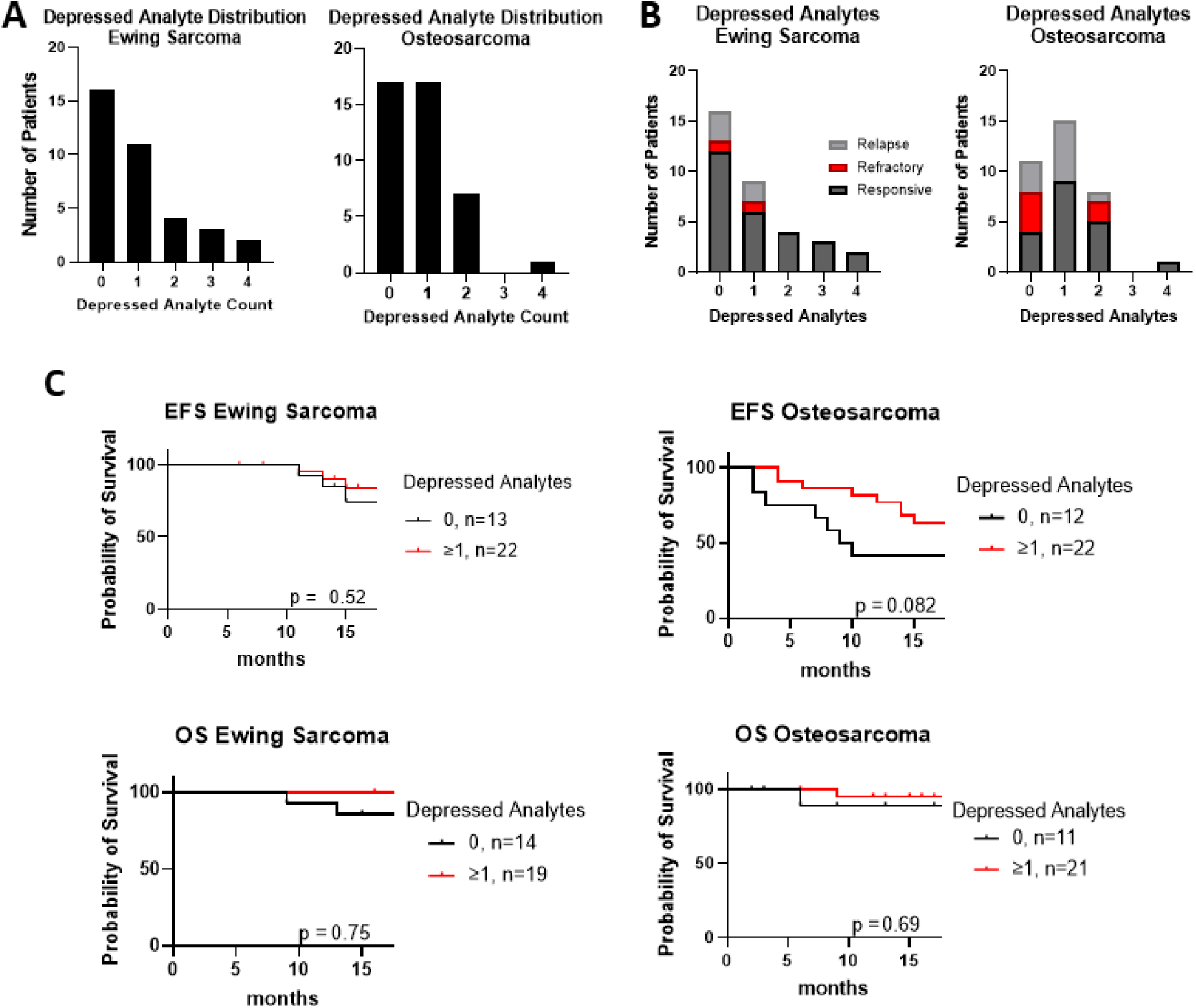
Comparison of outcome based on total number of depressed analytes, defined by bottom 5% of the healthy cohort’s mean. A) Distribution of osteosarcoma and Ewing sarcoma patients by their number of depressed cytokines. B) clinical outcomes for patients separated by how many depressed cytokines they have. C) Event free survival (EFS) and overall survival (OS) curves for Ewing sarcoma and osteosarcoma grouped based on number of depressed cytokines

The plasma levels of individual cytokines differ in patients with sarcoma compared to healthy controls. Levels of IL-1β, IL-4, IL-6, CXCL5, CXCL12, CXCL14, MIF, IGF-1, TGFβ-1, and CCL21 were increased in one or more sarcoma cohorts compared with controls, and levels of IL-15 and IL-16 were significantly lower in one or more sarcoma cohorts compared to healthy controls (Figure 4 and Supplementary Table 2). The remaining analytes, IL-8, IL-10, IL-12 P40, IL-24, IL-27, and CXCL10 were found in similar plasma concentrations in both patients with sarcoma and healthy subjects. ROC and AUC analyses of patients with bone sarcomas demonstrated that many of the cytokines with differential expression when compared to healthy controls have sufficiently significant sensitivity and specificity to be considered for use as part of clinical decision-making and monitoring (Supplementary Figure 2).

**Figure 4.**
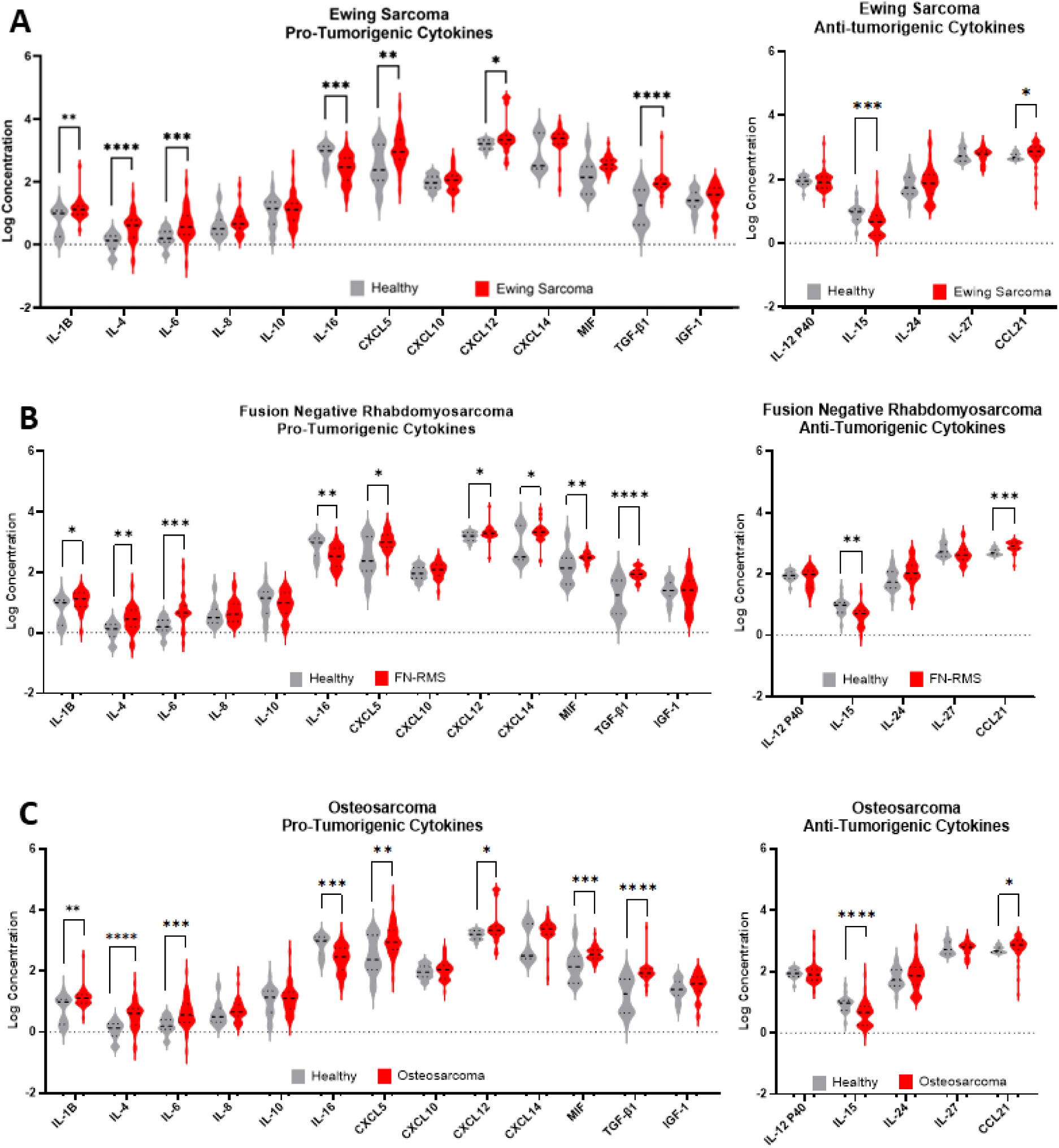
Pro-tumorigenic and anti-tumorigenic cytokine levels comparing healthy to A) Ewing sarcoma (ES), B) fusion negative rhabdomyosarcoma (FN-RMS) and C) osteosarcoma (OS). Statistical comparison using Mann-Whitney test. *, **, ***, and **** denote p<0.05, p<0.01, p<0.001, p<0.0001

Next, we tested whether median cytokine levels in patients with Ewing sarcoma, osteosarcoma, and patients in the “mixed cohorts” subgroup correlate with clinically relevant parameters such as age, stage at diagnosis, or clinical outcome. Adult patients in the “mixed cohorts” group had statistically significantly lower levels of IL-6 (0.29 pg/mL vs 0.64 pg/mL; p<0.05) and higher levels of MIF (2.67 pg/mL vs 2.49 pg/mL; p<0.05) than pediatric patients. Similarly, MIF levels were elevated in adult patients with Ewing sarcoma compared with pediatric patients (2.83 pg/mL vs 2.50 pg/mL; p<0.05). No cytokine levels varied by age in the osteosarcoma cohort. Interestingly, in the “mixed cohorts” patients, those who presented with metastatic disease had a higher median level of IL-1β than those presenting with localized disease (1.23 pg/mL vs. 1.10 pg/mL; p=0.022). Although this finding did not hold true for patients with Ewing sarcoma or osteosarcoma, in each of these cohorts there was a trend in that direction (Ewing sarcoma: 1.21 pg/mL vs. 1.07 pg/mL, p=0.06; osteosarcoma 1.32 pg/mL vs. 1.05 pg/mL, p=0.068). No other cytokine levels had a significant association with stage, and no median cytokine levels were associated with outcome. These and other non-significant values are displayed in Supplementary Table 3.

In contrast to comparisons based on median cytokine levels, high level expression (defined as the upper 25^th^ percentile of the cohort) of several cytokines correlated with 18-month OS and EFS (Figure 5 and Supplementary Table 4). Patients with osteosarcoma who have high IL-24 levels had poorer EFS (p=0.018) but slightly better OS (p=0.040). Patients with osteosarcoma showing high CXCL5 had poorer OS (p=0.010) but not EFS. Patients with Ewing sarcoma showing high levels of CXCL10 had slightly better OS (p=0.049), but IL-24 and CXCL5 levels had no prognostic significance. When analyzing all sarcomas unsegregated by diagnosis, we found that patients with high CXCL12 levels had shorter OS than those with low levels (p=0.0040).

**Figure 5.**
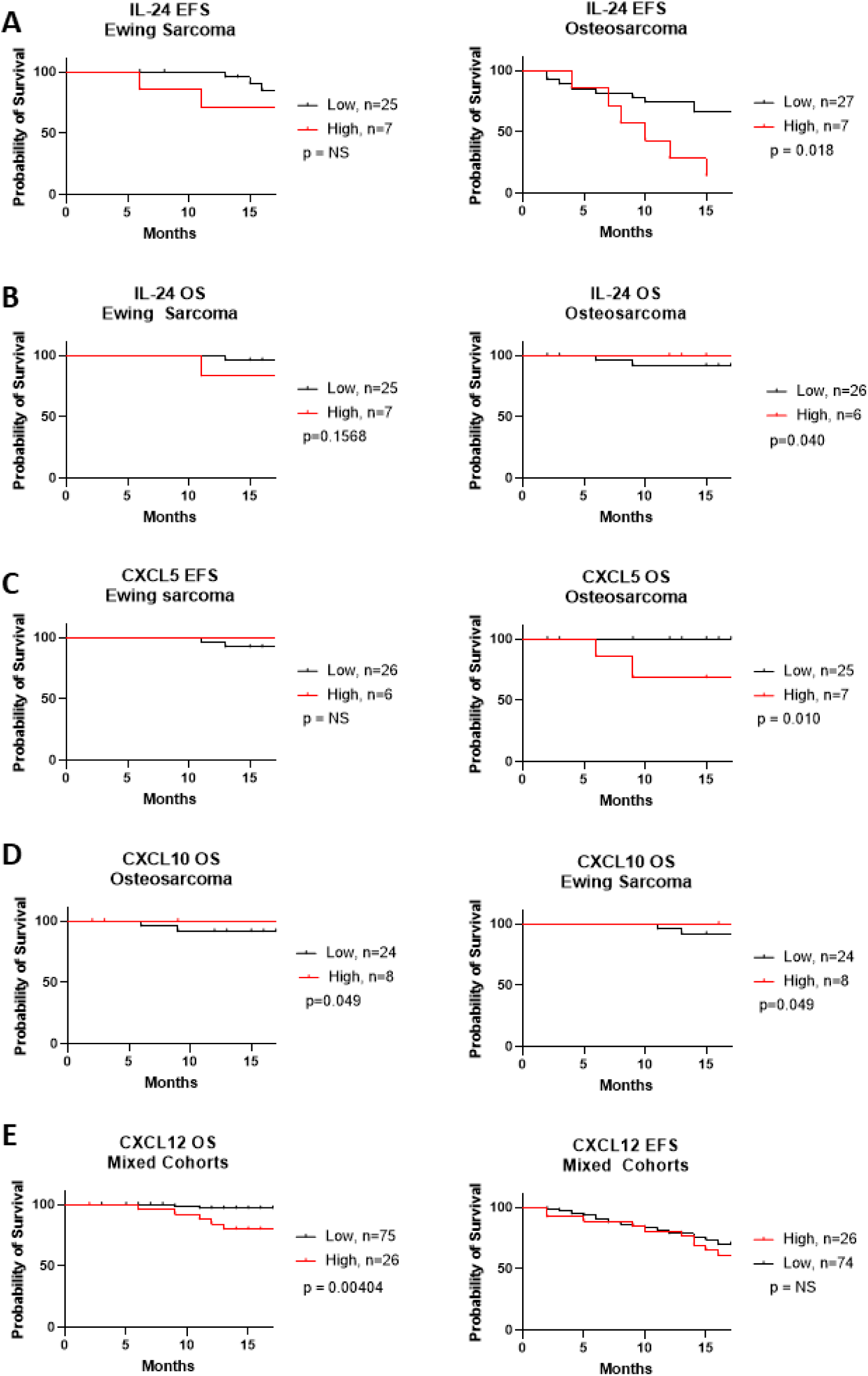
Cytokines that affected Event free survival (EFS) and overall survival (OS) of osteosarcoma and Ewing sarcoma. Significance determined by log rank test

Of the cytokines examined, IL-1β emerged as the cytokine with the most significant correspondence to features of disease progression. As mentioned above, IL-1β expression was significantly higher in the plasma of patients with metastatic disease from the mixed cohort (Figure 6A; p=0.022). Patients with Ewing sarcoma and osteosarcoma exhibited a similar trend but with subthreshold significance (Ewing sarcoma p=0.060; osteosarcoma p=0.068). Although 23.1% of patients with Ewing sarcoma who had low IL-1β had metastatic disease compared with 42.9% with high IL-1β (p=NS), in the osteosarcoma cohort only 4% of patients with low IL-1β had metastatic disease compared to 37.5% in the high IL-1β group (Figure 6B; p=0.036). In addition, patients with osteosarcoma who had high IL-1β levels had worse EFS than those with low IL-1β (p=0.0097). In contrast, high IL-1β did not predict worse EFS in patients with Ewing sarcoma (Figure 6C; p=0.36).

**Figure 6.**
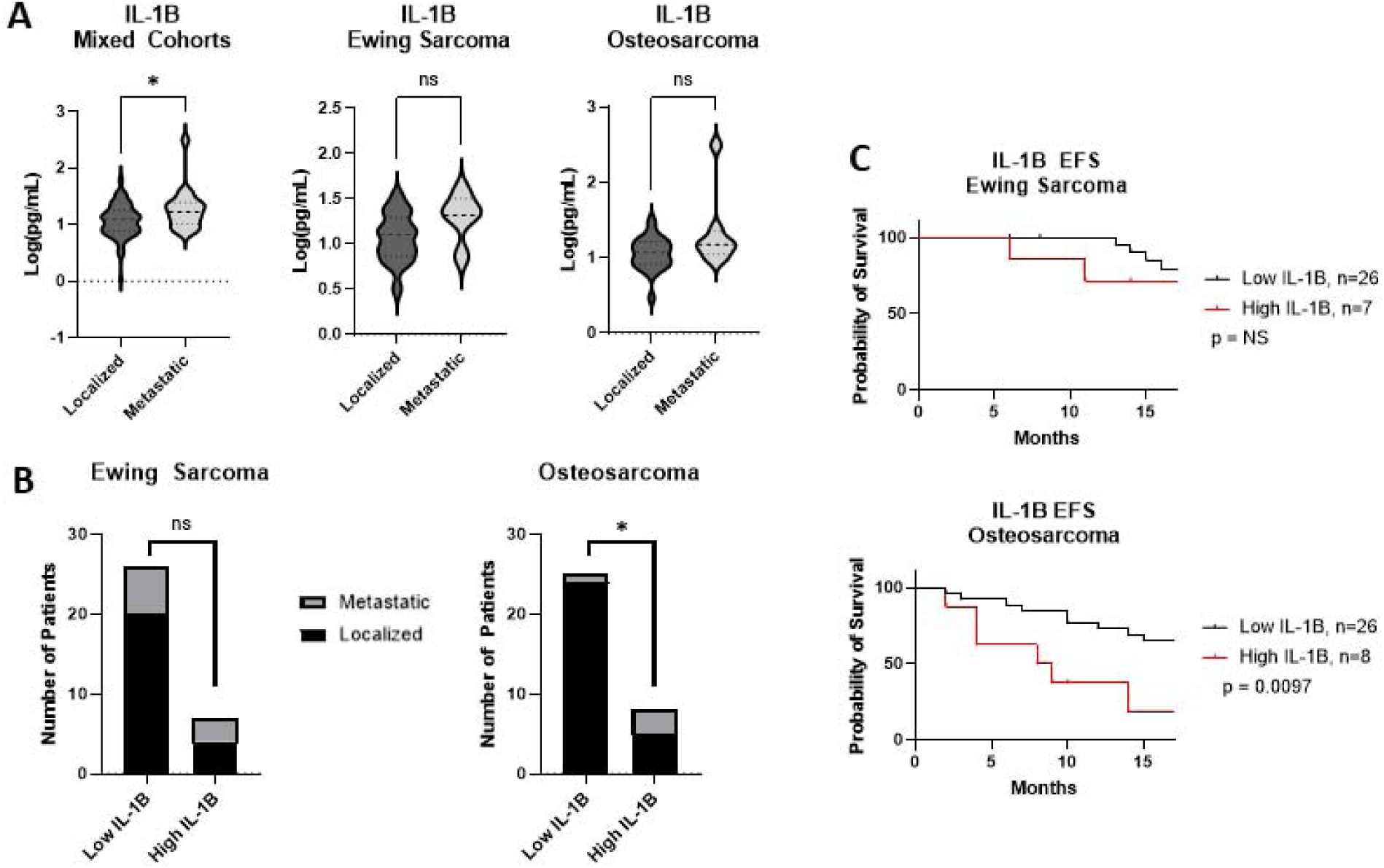
IL-1β is associated with worse outcomes in sarcomas. A) Mann-Whitney test comparing IL-1β in patients that had metastatic disease compared to localized in all patients, not separated by diagnosis, with Ewing sarcoma, and Osteosarcoma. B) Patients stratified by high (>25%) or low expression of IL-1β. Number of those patients that had metastatic or localized disease. Statistical analysis by Fisher’s exact test. C) Effect of IL-1β expression on Event free survival (EFS) of osteosarcoma and Ewing sarcoma. Significance determined by log rank test.

## Discussion

Sarcomas represent one of the most common pediatric cancers, and treatment options remain limited, especially for patients who present with metastatic disease^10^. This group of tumors is also unresponsive to standard cancer immunotherapies, such as immune checkpoint inhibitors (ICI)^43^. This study is the first to document the potential treatment-informative value of an 18-cytokine panel in the plasma of pediatric, adolescent, and young adult patients with sarcomas. It includes information on cytokines that have been poorly studied in sarcoma peripheral blood samples, such as IL-24, IL-27, CCL21, CXCL5, and CXCL14. It also identified one cytokine, IL-1β, that is associated with a targeted therapeutic and thus the potential for immediate evaluation in a clinical trial. We demonstrated that global cytokine profiling can distinguish healthy subjects from patients newly diagnosed with sarcoma. This difference is meaningful and non-random, as indicated by the finding of robust AUC results in ROC graphs as well as significant correlations between analyzed cytokines. Patients with different sarcoma diagnoses were not distinguishable from one another, indicating that this cytokine panel is insufficient to identify differences in the inflammatory profiles of different sarcoma subtypes, but does suggest that there may be shared immune processes active in the TME of different sarcomas.

Using unsupervised hierarchical clustering, three endotypes were apparent among patients. The group with the worst EFS and OS for osteosarcoma and Ewing sarcoma, Group 1, was defined by an increase in CXCL5, CXCL12, and MIF. These are protumorigenic chemokines are implicated in recruiting and activating myeloid-derived suppressor cells (MDSCs) and regulatory T cells (Tregs), immune cell populations that impair anti-tumor responses, as well as inducing angiogenesis and metastasis^16,17,44,45^. Group 2 had few visibly apparent abnormalities in cytokine expression except for some patients with increased CXCL10 expression or decreased IL-12p40. Group 3 had the longest EFS and had increased levels of both pro and anti-tumorigenic interleukins and/or decreased expression of CXCL12, IL-16, CXCL10, and CCL21. These findings may indicate specific coordinated effects of cytokines to orchestrate an anti-tumorigenic response. These endotypes probably reflect coordinate expression of cytokines with complementary effects on the immune system. Interestingly, we also found that patients with 5 or more analytes elevated in conjugation were more likely to have diminished EFS. Other studies have reported similar findings in sarcomas^31,46^. The clinical significance of these findings will be addressed in future work.

It is interesting to note that statistically significant correlations between cytokine expression and outcomes, such as EFS and OS, are predominantly observed in patients with osteosarcoma. Osteosarcoma appears to be the only sarcoma in children, adolescents, and young adults with a track record (albeit a weak one) of response to ICI. For example, in SARC028, a clinical trial of pembrolizumab in advanced sarcomas, 5% of osteosarcoma patients had a partial response and 30% had stable disease, whereas none of the patients with Ewing sarcoma responded^47^. There have been additional case reports of patients with osteosarcoma responding to similar agents^48^. This may be because ICI primarily addresses adaptive immune pathways, whereas many of the cytokines of interest that we have elucidated herein are produced by and signal to more than just adaptive immune cells. Indeed, mifamurtide, a fully synthetic derivative of immunostimulatory muramyl dipeptide (MDP) is approved by the European Medicines Agency for the treatment of high-grade resectable non-metastatic osteosarcoma after macroscopically complete surgical resection. MDP is a bacterial cell wall derivative that binds to NOD2, leading to downstream NF-κB and MAPK signaling in monocytes and macrophages, with subsequent production of pro-inflammatory cytokines such as TNF-α, IL-1β, and IL-6^49^. There are no comparable results in Ewing sarcoma, suggesting a fundamental difference between these bone sarcomas that is also reflected in our results.

Analysis of individual cytokine levels demonstrated robust differences in cytokine concentration in patients with sarcoma compared to healthy subjects. Furthermore, dramatic increases in expression were mostly observed in pro-tumorigenic cytokines (IL-1β, IL-4, IL-6, CXCL5, CXCL12, CXCL14 MIF, and TGF-β1), with little, if any, differences in the levels of anti-tumorigenic cytokines such as IL-12 p40, IL-24, and IL-27 when compared with healthy controls. IL-15, a potent activator of NK and T cell anti-tumor activity^23,50^, was underexpressed in sarcoma cohorts. CCL21 was overexpressed, and overexpression of this chemokine, frequently considered anti-tumorigenic, may reflect a normal, coordinated immune response, with simultaneous high expression of both pro- and anti-inflammatory signaling molecules, a finding also reported in other studies^7,39^. This panel, then, suggests that the immune system is creating an environment both supportive and permissive for tumor growth and metastasis. These findings are supported by those of other studies, strengthening their role in sarcoma^13,27,30,31,46,51–54^

Our findings related to IL-1β are particularly translationally relevant. Plasma levels of IL-1β were higher in patients with sarcoma than in healthy controls, and patients with osteosarcoma in particular with elevated IL-1β levels at diagnosis were more likely to present with metastatic disease and had a shortened EFS. For patients with osteosarcoma, metastasis is the strongest determinant for prognosis at diagnosis; patients with localized disease have a 70% five-year survival rate, which drops to <20% for patients who present with metastasis^10^. Unlike many of the cytokines we evaluated, there are mechanistic studies linking IL-1β directly to the process of metastasis. IL-1β is, in part, secreted by tumor-associated macrophages in osteosarcoma, where it has been proposed to promotes metastasis, an effect which was reversed by anakinra treatment^55^. A more recent study using preclinical models of osteosarcoma showed that pulmonary metastasis is initiated by a subpopulation of disseminated tumor cells that produce cytokines such as IL-6 and CXCL8 in response to lung-epithelium-derived IL-1α^56^. Although IL-1α and IL-1β are distinct cytokines, they use the same receptor, and nonredundancy appears to be primarily based on spatial restrictions of expression^57^. Because Reinecke et al. demonstrated that Anakinra, an IL-1 receptor antagonist, inhibits pulmonary metastases in their preclinical model^56^, our clinical data linking plasma IL-1β levels with metastasis and EFS (wherein metastasis is the predominant event) provide strong justification for potential clinical trials of IL-1 inhibition to prevent metastasis in patients with osteosarcoma.

In summary, our cytokine profiling of newly diagnosed patients with sarcoma uncovered several tantalizing findings worthy of further investigation. We found that plasma cytokine levels in patients newly diagnosed with sarcoma differ from those in healthy controls, but not from one sarcoma type to another. We also discovered three distinct endotypes within the sarcoma patient groups. Although these do not have statistically significant prognostic significance based on our current data, these data remain immature, and re-evaluation in the future may demonstrate prognostic or other significance. Interestingly, in comparing Ewing sarcoma and osteosarcoma, the populations with the largest representation in this study, we found correlations between certain cytokines and survival in patients with osteosarcoma but not in patients with Ewing sarcoma. It is notable that osteosarcoma is at times responsive to ICI, whereas Ewing sarcoma is not. This highlights the importance of looking beyond adaptive immune cells alone and considering which processes of cell-intrinsic, innate and/or adaptive immunity contribute to each tumor at each stage of its development. It remains to be seen whether the correlation between cytokines and response to immunotherapy is causal (ie., osteosarcoma is responsive because it elicits a more robust response from the patient’s immune system) or merely correlative. Perhaps most importantly, our work, in the context of preclinical studies from other groups, supports a contribution of IL-1-mediated inflammation to osteosarcoma metastasis and would justify a clinical trial of IL-1 blockade to prevent metastatic recurrence in these patients.

## Data Availability

All data produced in the present study are available upon reasonable request to the authors

## Acknowledgments

This work was supported, in part by a grant from the National Cancer Institute (5R01CA262802 to DML). We are grateful for support from the Einstein CFAR biomarker core (funded by grant 5P30AI124414-05) and the Montefiore Einstein Comprehensive Cancer Center support grant (2P30CA013330). Clinical trial MCC 20320 is supported by the National Pediatric Cancer Foundation, who also provided financial support for this study.

---

**Supplementary Figure 1.**
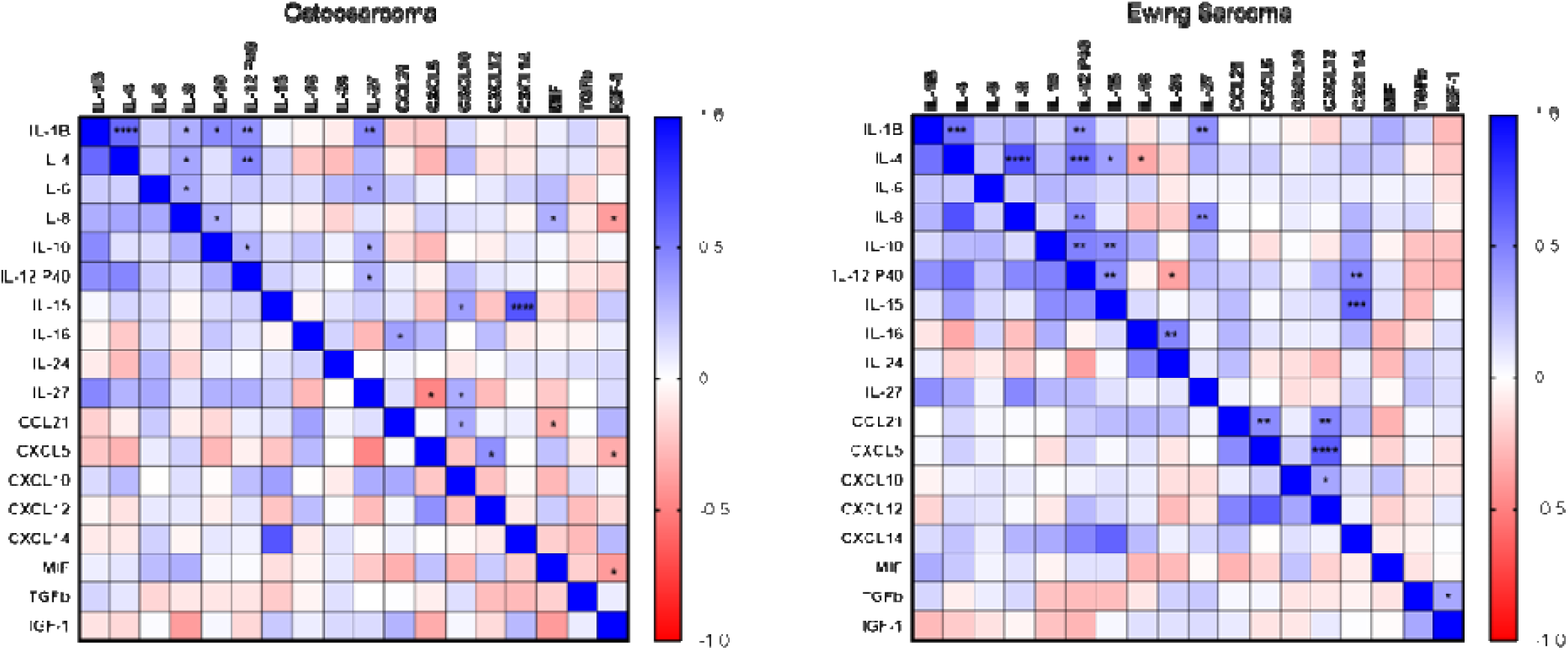
Correlation matrix of spearman correlation coefficients indicated by color gradient for each analyte in patients with (A) Osteosarcoma and (B) Ewing Sarcoma. Asterisks designate significance for that relationship where *, **, ***, and **** denote p<0.05, p<0.01, p<0.001, p<0.0001. Where no asterisks are shown, correlation was not statistically significant.

**Supplementary Figure 2.**
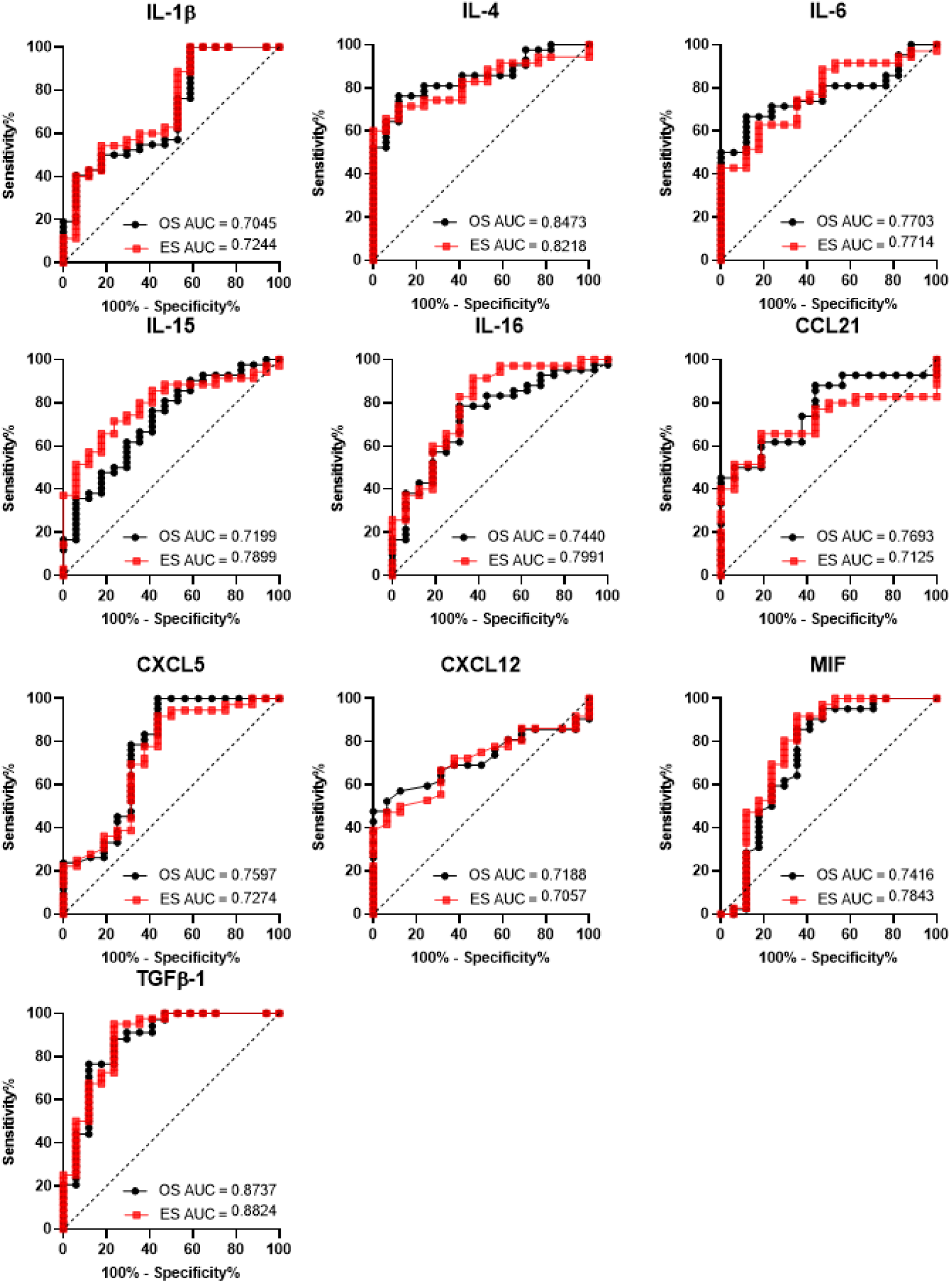
Receiver operating characteristics curves and area under the curve (AUC) for each cytokine that was significantly different in at least one sarcoma cohort compared to the healthy cohort. Curves compare healthy to osteosarcoma (OS) in black and Ewing sarcoma (ES) in red

**Supplementary Table 1:** Demographic characteristics of subjects included in this analysis.

| Cohort | Healthy<br>(n=17) | Ewing Sarcoma (n=36) | Fusion Positive<br>Rhabdomyosarcoma<br>(n=8) | Fusion Negative<br>Rhabdomyosarcoma<br>(n=24) | Osteosarcoma<br>(n=42) | Other Sarcoma (n=13) |
| --- | --- | --- | --- | --- | --- | --- |
| Age, n (%) |  |  |  |  |  |  |
| ≤21 | 1 (5.9%) | 29 (80.6%) | 8 (100%) | 22 (91.7%) | 38 (90.5%) | 9 (69.2%) |
| ≥22 | 16 (94.1%) | 7 (19.4%) | 0 (0%) | 2 (8.3%) | 4 (9.5%) | 4 (30.8%) |
| Gender, n (%) |  |  |  |  |  |  |
| Female | 7 (41.2%) | 18 (50.0%) | 6 (75.0%) | 11 (45.8%) | 20 (47.6%) | 6 (46.2%) |
| Male | 10 (58.8%) | 18 (50.0%) | 2 (25.0%) | 13 (54.2%) | 22 (52.4%) | 7 (53.8%) |
| Stage |  | n=34 | n=7 | n=23 | n=41 | n=12 |
| Localized |  | 25 (73.5%) | 6 (75.7%) | 21 (91.3%) | 36 (87.8%) | 9 (75.0%) |
| Metastatic |  | 11 (26.5%) | 1 (14.3%) | 2 (8.7%) | 5 (12.2%) | 4 (25.0%) |

**Supplementary Table 2:** Cytokine concentrations in plasma of healthy subjects and sarcoma patients at diagnosis. Median [IQR]. Statistical comparison of cohort to he using Mann-Whitney test where *, **, ***, and **** denote p<0.05, p<0.01, p<0.001, p<0.0001

| Analyte | IL-1β<br>pg/mL | IL-4<br>pg/mL | IL-6<br>pg/mL | IL-8<br>pg/mL | IL-10<br>pg/mL | IL-16<br>pg/mL | TGFβ-1<br>ng/mL | IGF-1<br>ng/mL |
| --- | --- | --- | --- | --- | --- | --- | --- | --- |
| Healthy | 0.99 [0.25-1.08] | 0.13 [-0.13-0.27] | 0.19 [0.08-0.42] | 0.5 [0.33-0.78] | 1.14 [0.65-1.35] | 2.99 [2.55-3.13] | 1.25 [0.63-1.74] | 1.4 [1.21-1.66] |
| Ewing sarcoma | 1.11 [0.96-1.23] | 0.61 [0.23-0.77] | 0.56 [0.32-0.92] | 0.66 [0.53-0.9] | 1.11 [0.77-1.32] | 2.47 [2.05-2.76] | 1.94 [1.82-2.08] | 1.58 [1.33-1.8] |
| *p* | ** | *** | *** | NS | NS | *** | **** | NS |
| Fusion positive<br>Rhabdomyosarcoma | 1.17 [0.79-1.62] | 0.76 [0.29-0.86] | 0.68 [-0.09-1.3] | 0.74 [0.58-1.05] | 1.02 [0.74-1.69] | 2.21 [1.84-2.78] | 1.8 [1.66-2.22] | 1.57 [1.24-1.92] |
| *p* | * | ** | NS | NS | NS | ** | * | NS |
| Fusion Negative<br>Rhabdomyosarcoma | 1.12 [0.87-1.33] | 0.46 [0.19-0.76] | 0.66 [0.53-0.9] | 0.62 [0.38-0.97] | 0.99 [0.64-1.33] | 2.53 [2.2-2.8] | 1.94 [1.83-2.06] | 1.41 [0.93-1.73] |
| *p* | * | ** | *** | NS | NS | ** | **** | NS |
| Osteosarcoma | 1.09 [0.88-1.32] | 0.5 [0.31-0.77] | 0.62 [0.29-0.95] | 0.7 [0.52-0.89] | 1 [0.73-1.33] | 2.45 [2.26-2.79] | 1.94 [1.79-2.09] | 1.7 [1.35-1.94] |
| *p* | ** | **** | *** | NS | NS | *** | **** | NS |
| Other Sarcoma | 1.08 [0.94-1.49] | 0.59 [0.27-0.85] | 0.56 [-0.2-0.66] | 0.62 [0.35-0.71] | 0.92 [0.76-1.5] | 2.53 [2.35-2.72] | 1.86 [1.73-1.94] | 1.61 [1.45-1.84] |
| *p* | * | *** | NS | NS | NS | * | ** | * |

| Analyte | CXCL5<br>pg/mL | CXCL10<br>pg/mL | CXCL12<br>pg/mL | CXCL14<br>Pg/mL | MIF<br>pg/mL |
| --- | --- | --- | --- | --- | --- |
| Healthy | 2.38 [2.04-3.18] | 1.97 [1.81-2.16] | 3.21 [3.05-3.33] | 2.51 [2.42-3.55] | 2.14 [1.61-2.48] |
| Ewing sarcoma | 2.95 [2.71-3.35] | 2.05 [1.75-2.2] | 3.34 [3.2-3.48] | 3.39 [3.2-3.52] | 2.55 [2.41-2.69] |
| *p* | ** | NS | * | NS | NS |
| Fusion positive<br>Rhabdomyosarcoma | 2.98 [2.74-3.26] | 2.08 [1.9-2.27] | 3.22 [2.96-3.28] | 3.29 [3.17-3.71] | 2.48 [2.46-2.79] |
| *p* | NS | NS | NS | NS | NS |
| Fusion Negative<br>Rhabdomyosarcoma | 3 [2.84-3.25] | 2.09 [1.9-2.26] | 3.29 [3.21-3.39] | 3.33 [3.22-3.47] | 2.49 [2.4-2.6] |
| *p* | * | NS | * | * | ** |
| Osteosarcoma | 3.03 [2.76-3.35] | 2.07 [1.94-2.27] | 3.37 [3.18-3.48] | 3.4 [3.27-3.47] | 2.5 [2.35-2.65] |
| *p* | ** | NS | * | NS | *** |
| Other Sarcoma | 2.86 [2.75-3.39] | 2 [1.92-2.11] | 3.36 [3.28-3.51] | 3.35 [3.27-3.66] | 2.61 [2.41-2.82] |
| *p* | * | NS | * | * | ** |

| Analyte | IL-12 P40<br>pg/mL | IL-15<br>pg/mL | IL-24<br>pg/mL | IL-27<br>pg/mL | CCL21<br>pg/mL |
| --- | --- | --- | --- | --- | --- |
| Healthy | 1.95 [1.84-2.06] | 0.98 [0.74-1.06] | 1.73 [1.54-2.07] | 2.72 [2.57-2.97] | 2.68 [2.63-2.78] |
| Ewing sarcoma | 1.9 [1.73-2.08] | 0.66 [0.25-0.87] | 1.87 [1.55-2.15] | 2.79 [2.6-2.88] | 2.87 [2.73-3.01] |
| *p* | NS | *** | NS | NS | * |
| Fusion positive<br>Rhabdomyosarcoma | 2.01 [1.85-2.08] | 0.81 [0.55-1.25] | 1.91 [1.53-2.13] | 2.69 [2.62-2.99] | 2.8 [2.62-2.9] |
| *p* | NS | NS | NS | NS | NS |
| Fusion Negative<br>Rhabdomyosarcoma | 1.98 [1.57-2.1] | 0.7 [0.39-0.81] | 2.03 [1.84-2.28] | 2.61 [2.46-2.79] | 2.91 [2.81-3.02] |
| *p* | NS | ** | NS | NS | *** |
| Osteosarcoma | 1.97 [1.85-2.09] | 0.76 [0.56-0.94] | 1.87 [1.58-2.08] | 2.64 [2.53-2.75] | 2.89 [2.75-2.97] |
| *p* | NS | **** | NS | NS | * |
| Other Sarcoma | 1.83 [1.55-2.12] | 0.79 [0.5-1.07] | 2.06 [1.56-2.39] | 2.74 [2.49-2.87] | 2.84 [2.77-2.96] |
| *p* | NS | NS | NS | NS | * |

**Supplementary Table 3.** Association Between Cytokine Level and Clinicopathological Parameters. Median values displayed with significance determined by Mann-Whitney test. Hazard ratio of high to low group for overall survival (OS) and event free survival (EFS) by log rank analysis. Significance marked on higher value where *, **, ***, and **** denote p<0.05, p<0.01, p<0.001, p<0.0001

| Clinicopathological Parameter | | IL-1β<br>pg/mL | IL-4<br>pg/mL | IL-6<br>pg/mL | IL-8<br>pg/mL | IL-10<br>pg/mL | IL-16<br>pg/mL | CXCL5<br>pg/mL | CXCL10<br>pg/mL | CXCL12<br>pg/mL | CXCL14<br>pg/mL | MIF<br>pg/mL | TGFβ-1<br>ng/mL | IGF-1<br>ng/mL |
| --- | --- | --- | --- | --- | --- | --- | --- | --- | --- | --- | --- | --- | --- | --- |
| Age |  |  |  |  |  |  |  |  |  |  |  |  |  |  |
| Mixed Cohorts | $\leq 21$ | 1.10 | 0.51 | 0.64 | 0.67 | 1.06 | 2.68 | 3.01 | 1.92 | 3.33 | 3.37 | 2.49 | 1.93 | 1.61 |
| | $\geq 22$ | 1.11 | 0.66 | 0.29 | 0.62 | 0.90 | 2.75 | 2.88 | 1.83 | 3.35 | 3.39 | 2.67 | 1.95 | 1.58 |
|  | *p* | NS | NS | * | NS | NS | NS | NS | NS | NS | NS | * | NS | NS |
| Ewing sarcoma |  |  |  |  |  |  |  |  |  |  |  |  |  |  |
| | $\leq 21$ | 1.13 | 0.51 | 0.58 | 0.65 | 1.13 | 2.53 | 2.95 | 2.04 | 3.34 | 3.40 | 2.50 | 1.92 | 1.58 |
| | $\geq 22$ | 1.04 | 0.64 | 0.28 | 0.84 | 0.91 | 2.33 | 3.28 | 2.10 | 3.36 | 3.40 | 2.83 | 2.03 | 1.70 |
|  | *p* | NS | NS | NS | NS | NS | NS | NS | NS | NS | NS | * | NS | NS |
| Osteo-sarcoma |  |  |  |  |  |  |  |  |  |  |  |  |  |  |
| | $\leq 21$ | 1.02 | 0.45 | 0.63 | 0.71 | 1.00 | 2.47 | 3.05 | 2.07 | 3.38 | 3.40 | 2.50 | 1.94 | 1.73 |
| | $\geq 22$ | 1.21 | 0.70 | 0.14 | 0.59 | 1.17 | 2.23 | 2.74 | 2.15 | 3.27 | 3.41 | 2.46 | 1.94 | 1.51 |
|  | *p* | NS | NS | NS | NS | NS | NS | NS | NS | NS | NS | NS | NS | NS |
| Stage |  |  |  |  |  |  |  |  |  |  |  |  |  |  |
| Mixed Cohorts | Localized | 1.10 | 0.54 | 0.59 | 0.68 | 0.99 | 2.67 | 2.99 | 1.96 | 3.33 | 2.50 | 3.38 | 1.93 | 1.62 |
|  | Metastatic | 1.23 | 0.66 | 0.63 | 0.71 | 1.10 | 2.75 | 2.89 | 1.81 | 3.29 | 2.53 | 3.39 | 1.93 | 1.50 |
|  | *p* | 0.022 | 0.26 | 0.51 | 0.94 | 0.34 | 0.36 | 0.86 | 0.58 | 0.27 | 0.92 | 0.20 | 0.55 | 0.36 |
| Ewing sarcoma |  |  |  |  |  |  |  |  |  |  |  |  |  |  |
|  | Localized | 1.07 | 0.60 | 0.49 | 0.65 | 1.12 | 2.46 | 2.95 | 2.00 | 3.33 | 3.41 | 2.54 | 1.94 | 1.68 |
|  | Metastatic | 1.21 | 0.69 | 0.75 | 0.76 | 1.02 | 2.51 | 3.42 | 2.06 | 3.38 | 3.40 | 2.50 | 1.93 | 1.33 |
|  | *p* | 0.060 | 0.36 | 0.20 | 0.49 | 0.65 | 0.36 | 0.26 | 0.58 | 0.90 | 0.96 | 0.70 | 0.59 | 0.061 |
| Osteo-sarcoma |  |  |  |  |  |  |  |  |  |  |  |  |  |  |
|  | Localized | 1.05 | 0.54 | 0.62 | 0.70 | 0.96 | 2.43 | 3.05 | 2.07 | 3.40 | 3.40 | 2.50 | 1.93 | 1.70 |
|  | Metastatic | 1.32 | 0.48 | 0.60 | 0.83 | 1.25 | 2.45 | 2.88 | 2.26 | 3.20 | 3.40 | 2.45 | 2.04 | 1.98 |
|  | *p* | 0.068 | 0.65 | 0.74 | 0.77 | 0.30 | 0.59 | 0.33 | 0.20 | 0.11 | 0.58 | 0.90 | 0.58 | 0.16 |

Supplementary Table 3. Association Between Cytokine Level and Clinicopathological Parameters.
| Clinicopathological Parameter |  | IL-12p40<br>pg/mL | IL-15<br>pg/mL | IL-24<br>pg/mL | IL-27<br>pg/mL | CCL21<br>pg/mL |
| --- | --- | --- | --- | --- | --- | --- |
| Age |  |  |  |  |  |  |
| Mixed Cohorts | $\leq 21$ | 1.97 | 0.69 | 2.07 | 2.88 | 2.52 |
| | $\geq 22$ | 1.84 | 0.80 | 2.06 | 2.77 | 2.41 |
|  | *p* | NS | NS | NS | NS | NS |
| Ewing sarcoma |  |  |  |  |  |  |
| | $\leq 21$ | 1.84 | 0.65 | 1.97 | 2.78 | 2.90 |
| | $\geq 22$ | 1.98 | 0.67 | 1.81 | 2.85 | 2.83 |
|  | *p* | NS | NS | NS | NS | NS |
| Osteo-sarcoma |  |  |  |  |  |  |
| | $\leq 21$ | 1.97 | 0.73 | 1.89 | 2.63 | 2.93 |
| | $\geq 22$ | 2.00 | 0.91 | 1.64 | 2.79 | 2.75 |
|  | *p* | NS | NS | NS | NS | ** |
| Stage |  |  |  |  |  |  |
| Mixed Cohorts | Localized | 1.96 | 0.70 | 2.06 | 2.89 | 2.48 |
|  | Metastatic | 1.96 | 0.75 | 2.06 | 2.84 | 2.57 |
|  | *p* | 0.56 | 0.41 | 0.35 | 0.16 | 0.44 |
| Ewing sarcoma |  |  |  |  |  |  |
|  | Localized | 1.84 | 0.65 | 1.97 | 2.78 | 2.90 |
|  | Metastatic | 1.98 | 0.67 | 1.81 | 2.75 | 2.84 |
|  | *p* | 0.18 | 0.98 | 0.37 | 0.17 | 0.31 |
| Osteo-sarcoma |  |  |  |  |  |  |
|  | Localized | 1.97 | 0.73 | 1.86 | 2.63 | 2.89 |
|  | Metastatic | 2.08 | 0.89 | 1.99 | 2.75 | 2.96 |
|  | *p* | 0.38 | 0.19 | 0.71 | 0.15 | 0.49 |

**Supplementary Table 4.**
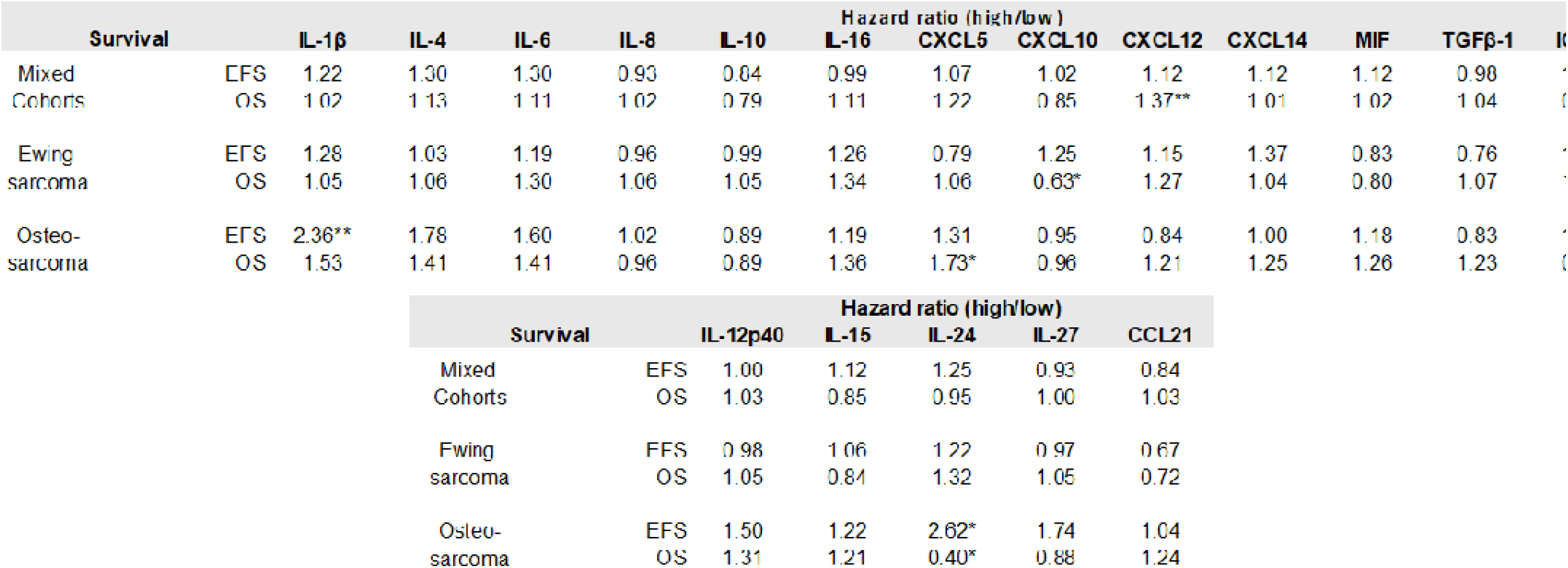
Hazard ratio of high to low group for overall survival (OS) and event free survival (EFS) by log rank analysis. Significance marked such that *, **, ***, and **** denote p<0.05, p<0.01, p<0.001, p<0.0001

## Notes

### Competing Interest Statement

The authors have declared no competing interest.

### Funding Statement

This study was funded by the National Pediatric Cancer Foundation

### Author Declarations

Research subjects were enrolled in an ongoing multi-center clinical trial, MCC20320. This clinical trial was approved by Advarra, and was conducted in accordance with the Ethical Principles and Guidelines for the Protection of Human Subjects of Research.

